# Humoral Immunity Against Orthopoxvirus in Vaccinated and Unvaccinated Individuals during 2022 Mpox Outbreak

**DOI:** 10.1101/2025.03.16.25324075

**Authors:** Yijia Li, Michael B. Townsend, Shanshan Li, Quinn E Testa, Tom Medvec, Elizabeth A. Thompson, Frank J. Palella, Matthew J. Mimiaga, James B. Brock, Maria L. Alcaide, Anandi N. Sheth, Michelle Floris-Moore, Aruna Chandran, Audrey L. French, Phyllis C. Tien, Daniel J Merenstein, Michael Augenbraun, Anjali Sharma, Caitlin A. Moran, Charles R. Rinaldo, Bernard JC. Macatangay, Panayampalli S. Satheshkumar, Ken S. Ho

## Abstract

Little is known about serological responses to MVA-BN (JYNNEOS) against mpox in elderly individuals with or without HIV. In this study, MVA-BN induced sustained IgG levels regardless of HIV status even up to one year. Birth before 1973 correlated with higher IgG. MVA-BN unvaccinated individuals with HIV had lower IgG than vaccinated.

---

Monkeypox virus (MPXV) is an orthopoxvirus that causes mpox, a clinical illness similar to smallpox. In 2022, a global outbreak of mpox was caused by Clade IIb MPXV and disproportionately affected men who reported having sex with men. As of December 31, 2024, a total of 34,490 cases of mpox were reported to the United States (US) Centers for Disease Control and Prevention (CDC)^1^. Even though most cases were mild and self-limited, severe or even fatal cases can be seen in people with HIV (PWH) ^2^. Routine childhood smallpox vaccination was terminated in 1972 after the disease was eradicated in the US^3^, resulting in few civilians having received smallpox vaccinations until the 2022 outbreak when the Modified Vaccinia Ankara-Bavarian Nordic (MVA-BN) vaccine (JYNNEOS) became available. Although exact correlates of protection following MVA-BN are not well-characterized, MVA-BN protects against mpox^4^. Breakthrough infections after MVA-BN are rare; in most cases, the symptoms were milder^5,6^. Few studies have focused on the long-term immunogenicity and effectiveness in PWH. Prior efforts found in the short term (one month after immunizations), MVA-BN is immunogenic but HIV infection appears to negatively affect neutralizing antibody response^7^. However, existing data on the long-term immune response to MVA-BN raise concerns, as one study has reported that MPXV-IgG levels after either one or two doses of MVA-BN were reported to decline almost to pre-vaccination levels 12 months after immunizations^8^. In that study, however, most participants were young, HIV-negative (only 9% had HIV), and otherwise did not have previous smallpox vaccination exposure ^8^. Little is known about the association between HIV, previous smallpox vaccination, and long-term vaccine immunogenicity in regard to aging populations. To this end, we leverage Multicenter AIDS Cohort Study (MACS)/Women’s Interagency HIV Study (WIHS) Combined Cohort Study (MWCCS), a multicenter prospective cohort study of men and women with or at risk for HIV, to evaluate serological response to either historic smallpox vaccinations or MVA-BN vaccinations during 2022 outbreaks.

Of the 114 MWCCS participants included in the study (median age of 64 years; Table S1), 75% were male and 89% were born before 1973, and 52 (46%) were HIV seropositive, with an HIV-1 viral suppression rate of 88.5% (HIV-1 viral load<50 copies/ml). In 20 participants receiving MVA-BN, 15 (75%) received two doses. In the analysis of results from all participants, the MVA-BN vaccinated group showed higher anti-orthopoxvirus IgG levels at Timepoint 3 (6 months after the first dose of MVA-BN or after 6 months of the 2022 outbreak, adjusted P<0.0001, Figure 1; Table S2) relative to participants that had not received MVA-BN. Longitudinally, the MVA-BN vaccinated group showed a persistent increase in IgG levels. The number of vaccine doses received and HIV serostatus were not associated with IgG levels, while birth before 1973, or those who likely had childhood smallpox vaccination, was significantly associated with higher IgG levels (Figure S1; Table S3). IgG levels were stably low in those who had not received MVA-BN. HIV seropositivity was negatively associated with IgG levels while birth before 1973 was again significantly associated with higher IgG levels (Figure S2; Table S4). No participants reported ever having a positive mpox test result.

**Figure 1.**
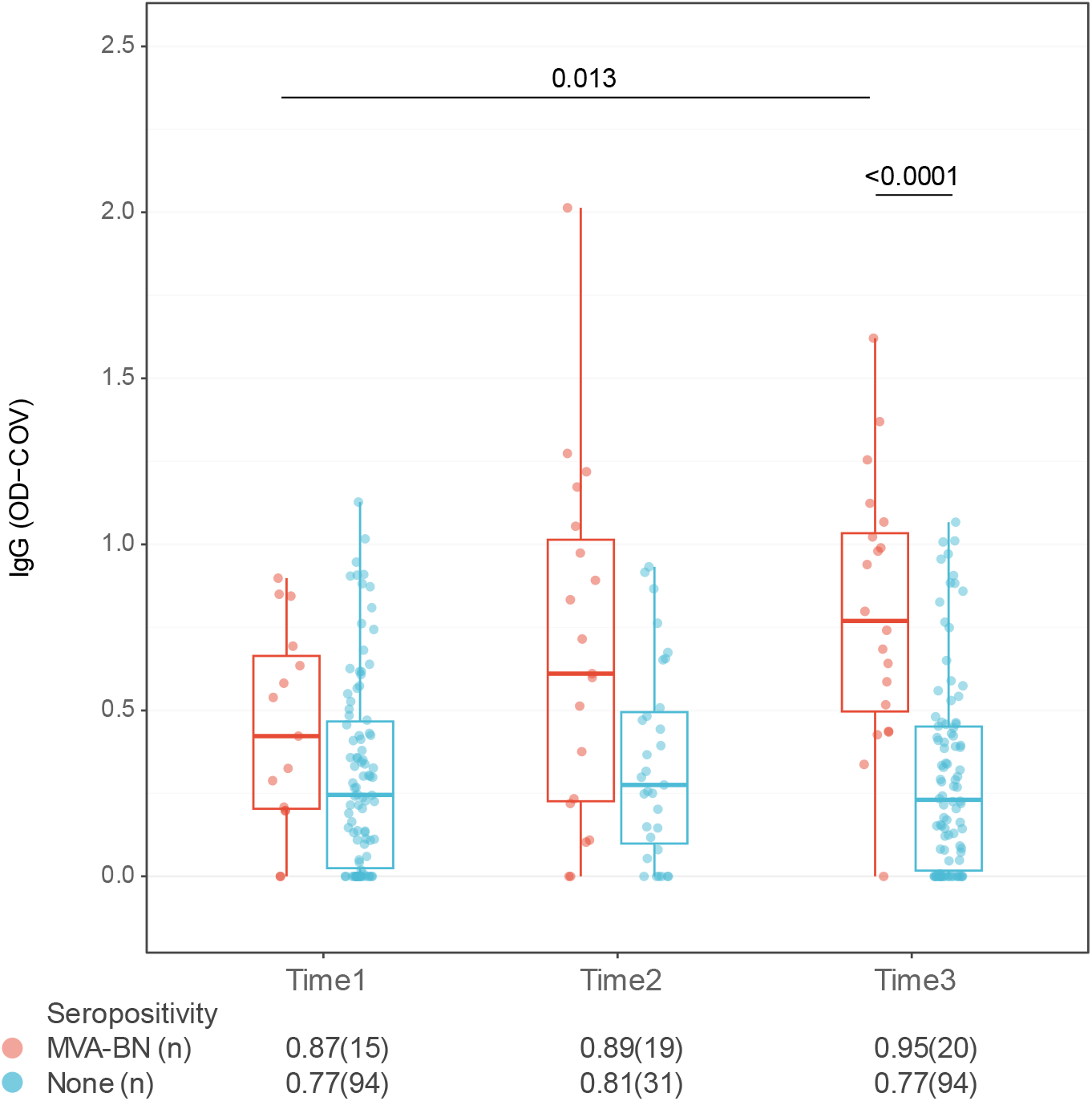
Anti-orthopoxvirus IgG levels in MVA-BN vaccinated (MVA-BN) and those without MVA-BN vaccination (None) groups. Time 1: Before first dose of vaccinations (vaccinated group) or before 2022 outbreak (unvaccinated groups). Time 2: < 6 months of first dose of vaccinations (vaccinated group) or within 6 months of 2022 outbreak (designated as June 1st, 2022). Time 3: ≥6 months after first dose of MVA-BN vaccinations (vaccinated group) or over 6 months since 2022 outbreak. P values between groups were calculated using Wilcoxon rank sum test and P values within groups were calculated using Wilcoxon singed rank test. Final P values were adjusted for multiple comparisons using the Benjamini-Hochberg method. Nonsignificant P values were not shown. Details on time interval between each time point and vaccinations were shown in Table S2 (appendix p3).

Our results show that among older PWH who probably received childhood smallpox vaccination using a live, replicating vaccine (e.g., Dryvax), there is a detectable level of humoral immunity against mpox. While there is no clear correlate of protection against mpox, as antibody levels from childhood vaccination wane over time, there may be value in providing at least one dose of MVA-BN to these individuals as they may have insufficient protection. Although limited by a small sample size and only having measured binding IgG levels to whole orthopoxvirus antigens rather than neutralization, our study is the first to enroll some of the most vulnerable participants (elderly individuals with HIV).

In summary, we herein demonstrate MVA-BN provides comparable humoral immunogenicity regardless of HIV serostatus and significantly increases anti-orthopoxvirus IgG levels that are sustained even 6 to 12 months post-vaccination. In MVA-BN-unvaccinated PWH who previously received smallpox vaccinations, antibody levels were lower than among persons without HIV. These findings may guide vaccination recommendations for those living with HIV, and especially for those of older age.

## Funding sources and conflict of interests

Data in this manuscript were collected by the MACS/WIHS Combined Cohort Study (MWCCS). The contents of this publication are solely the responsibility of the authors and do not represent the official views of the National Institutes of Health (NIH) or the Centers for Disease Control and Prevention (CDC). MWCCS (Principal Investigators): Atlanta CRS (Ighovwerha Ofotokun, Anandi Sheth, and Gina Wingood), U01-HL146241; Baltimore CRS (Todd Brown and Joseph Margolick), U01-HL146201; Bronx CRS (Kathryn Anastos, David Hanna, and Anjali Sharma), U01-HL146204; Brooklyn CRS (Deborah Gustafson and Tracey Wilson), U01-HL146202; Data Analysis and Coordination Center (Gypsyamber D’Souza, Stephen Gange and Elizabeth Topper), U01-HL146193; Chicago-Cook County CRS (Mardge Cohen, Audrey French, and Ryan Ross), U01-HL146245; Chicago-Northwestern CRS (Steven Wolinsky, Frank Palella, and Valentina Stosor), U01-HL146240; Northern California CRS (Bradley Aouizerat, Jennifer Price, and Phyllis Tien), U01-HL146242; Los Angeles CRS (Roger Detels and Matthew Mimiaga), U01-HL146333; Metropolitan Washington CRS (Seble Kassaye and Daniel Merenstein), U01- HL146205; Miami CRS (Maria Alcaide, Margaret Fischl, and Deborah Jones), U01-HL146203; Pittsburgh CRS (Jeremy Martinson and Charles Rinaldo), U01-HL146208; UAB-MS CRS (Mirjam-Colette Kempf, James B. Brock, Emily Levitan, and Deborah Konkle-Parker), U01- HL146192; UNC CRS (M. Bradley Drummond and Michelle Floris-Moore), U01-HL146194. The MWCCS is funded primarily by the National Heart, Lung, and Blood Institute (NHLBI), with additional co-funding from the Eunice Kennedy Shriver National Institute Of Child Health & Human Development (NICHD), National Institute On Aging (NIA), National Institute Of Dental & Craniofacial Research (NIDCR), National Institute Of Allergy And Infectious Diseases (NIAID), National Institute Of Neurological Disorders And Stroke (NINDS), National Institute Of Mental Health (NIMH), National Institute On Drug Abuse (NIDA), National Institute Of Nursing Research (NINR), National Cancer Institute (NCI), National Institute on Alcohol Abuse and Alcoholism (NIAAA), National Institute on Deafness and Other Communication Disorders (NIDCD), National Institute of Diabetes and Digestive and Kidney Diseases (NIDDK), National Institute on Minority Health and Health Disparities (NIMHD), and in coordination and alignment with the research priorities of the National Institutes of Health, Office of AIDS Research (OAR). MWCCS data collection is also supported by UL1-TR000004 (UCSF CTSA), UL1-TR003098 (JHU ICTR), UL1-TR001881 (UCLA CTSI), P30-AI-050409 (Atlanta CFAR), P30-AI-073961 (Miami CFAR), P30-AI-050410 (UNC CFAR), P30-AI-027767 (UAB CFAR), P30-MH-116867 (Miami CHARM), UL1-TR001409 (DC CTSA), KL2-TR001432 (DC CTSA), and TL1-TR001431 (DC CTSA). The authors declared no other conflicts of interest pertinent to this study.

The authors gratefully acknowledge the contributions of the study participants and the dedication of the staff at the MWCCS sites. We also thank Dr. Carolyn Williams from the National Institute of Allergy and Infectious Diseases, National Institutes of Health for her insightful comments.

## Data Availability

All data produced in the present work are contained in the manuscript.

