## Supplementary Materials for "Humoral Immunity Against Orthopoxvirus in Vaccinated and Unvaccinated Individuals during 2022 Mpox Outbreak"

### Supplementary Methods

Participants

Participants enrolled in the MWCCS cohort who had self-reported mpox and smallpox vaccination information were included in this study. Additional inclusion criteria were:

(1) For participants vaccinated with MVA-BN, available serum samples from pre-vaccination, less than six months after final dose, and over six months after final dose.
(2) For participants not vaccinated with MVA-BN during this outbreak, available serum samples from pre-outbreak (before 2022), 6 months within outbreak, and over 6 months after outbreak started.
(3) All sexes.
(4) Age≥18 years.
(5) Available HIV-related information.

Serum samples were obtained at each MWCC scheduled visits and cryopreserved at central repository at Johns Hopkins University Data Analysis and Coordination Center (DACC).

Anti-Orthopoxvirus binding IgG measurement

Anti-Orthopoxvirus binding IgG measurement was reported in our previous studies. Briefly, serum samples were tested for Orthopoxvirus IgG at 1:100 dilution using MVA-BN as antigen. We used Optical Density (OD) - cutoff value (COV) (OD-COV) derived from ELISA to determine anti-Orthopoxvirus IgG levels [1,2]. Results below 0 were censored to 0 and considered seronegative.

Statistical analysis

Categorical variables were summarized using number and percentage and compared using either Chi-Squared test or Fisher Exact test. Continuous variables were summarized using median and interquartile range and compared using nonparametric methods. For two independent groups, we used Wilcoxon Rank Sum test and for paired samples (longitudinal measurements) we used Wilcoxon Signed Rank test. P values from multiple comparisons were adjusted using the Benjamin-Hochberg method. For longitudinal repeated measurements, we used Generalized Estimating Equation (GEE) to estimate variables associated with IgG levels using autoregressive correlation structure using “geepack” based on R platform (version 1.3.12). In longitudinal GEE analysis, timepoints earlier than 12 months before final vaccinations (vaccinated group) or June 1st, 2022 (unvaccinated group), were censored to -12 months. In addition, in the longitudinal GEE analysis, if the second serum sampling time point fell between two vaccination doses, this participant will be labeled as receiving one dose of MVA-BN for the first two time points and labeled as one dose at the third time point. In addition, the time variable for the first 2 time points will be counted as the interval between sampling and the first dose of vaccination, and the third time point will be counted as the interval between the second dose of vaccination and the sampling time point. All statistical analyses were performed using R (version 4.3.1).

**Supplementary Tables**

**Table S1**. Demographic information.

| Characteristic | Overall N = 114 | MVA-BN N = 20 | None N = 94 | P |
|---|---|---|---|---|
| Age, (IQR) | 64 (58, 69) | 64 (61, 67) | 64 (58, 69) | 0.8 |
| Born before 1973 | | | | >0.9 |
| No | 12 (10.5%) | 2 (10.0%) | 10 (10.6%) | |
| Yes | 102 (89.5%) | 18 (90.0%) | 84 (89.4%) | |
| HIV Status | | | | >0.9 |
| Negative | 62 (54.4%) | 11 (55.0%) | 51 (54.3%) | |
| Positive | 52 (45.6%) | 9 (45.0%) | 43 (45.7%) | |
| Sex | | | | 0.003 |
| Female | 28 (24.6%) | 0 (0.0%) | 28 (29.8%) | |
| Male | 86 (75.4%) | 20 (100.0%) | 66 (70.2%) | |
| Race | | | | 0.8 |
| American or Alaskan Native | 1 (0.9%) | 0 (0.0%) | 1 (1.1%) | |
| Asian | 1 (0.9%) | 0 (0.0%) | 1 (1.1%) | |
| Black/African-American | 24 (21.1%) | 3 (15.0%) | 21 (22.3%) | |
| White/Caucasian | 85 (74.6%) | 17 (85.0%) | 68 (72.3%) | |
| Other | 2 (1.8%) | 0 (0.0%) | 2 (2.1%) | |
| Multi-Racial | 1 (0.9%) | 0 (0.0%) | 1 (1.1%) | |
| Hispanic ethnicity | 14 (12.3%) | 2 (10.0%) | 12 (12.8%) | >0.9 |
| Number of MVA-BN doses | | | | |
| 1 | | 5 (25.0%) | | |
| 2 | | 15 (75.0%) | | |
| CD4 cell count, cells/mm$^3$ (IQR) * | 726 (531, 970) | 698 (640, 800) | 763 (523, 988) | 0.5 |
| Nadir CD4 cell count, cells/mm$^3$ (IQR) * | 235 (127, 363) | 298 (240, 391) | 215 (111, 356) | 0.093 |
| Viral suppression<50 copies/ml * | 46 (88.5%) | 8 (88.9%) | 38 (88.4%) | >0.9 |

MVN-BN, Modified Vaccinia Ankara-Bavarian Nordic (MVA-BN) vaccine; IQR, interquartile range. *, only applicable to people with HIV.

**Table S2.** Time interval between vaccinations and each time point shown in Figure 1.

| Group | Beginning time point | Time 1 (months) | Time 2 (months) | Time 3 (months) |
|---|---|---|---|---|
| MVA-BN | First dose MVA-BN | -11 (-28 - -9.5) | 1 (0 - 2) | 11.5 (8 - 13) |
| MVA-BN | Last dose MVA-BN | -13 (-28.5 - -11) | 0 (0 - 0.5) | 9.5 (6 - 12) |
| Unvaccinated | June 1st, 2022 | -11 (-13 - -9) | 1 (0 - 2) | 11 (10 - 13) |

**Table S3**. GEE coefficients for the MVA-BN vaccinated group. All vaccinated participants were male assigned at birth.

| Variable | Estimate | 95% CI | P |
|---|---|---|---|
| Time (months) | 0.009 | 0 - 0.01 | 0.001 |
| HIV seropositive (seronegative as reference) | -0.10 | -0.39 - 0.18 | 0.5 |
| Two doses of MVA-BN (one dose as reference) | 0.19 | -0.03 - 0.41 | 0.1 |
| Born before 1973 (after 1973 as reference) | 0.62 | 0.3 - 0.94 | <0.001 |

**Table S4**. GEE coefficients for the MVA-BN unvaccinated group.

| Variable | Estimate | 95% CI | P |
|---|---|---|---|
| Time (months) | 0 | 0 - 0 | 0.4 |
| HIV seropositive (seronegative as reference) | -0.14 | -0.24 - -0.04 | 0.008 |
| Born before 1973 (after 1973 as reference) | 0.29 | 0.21 - 0.37 | <0.001 |
| Male sex (Female as reference) | 0.051 | -0.06 - 0.16 | 0.4 |

### Supplementary Figures

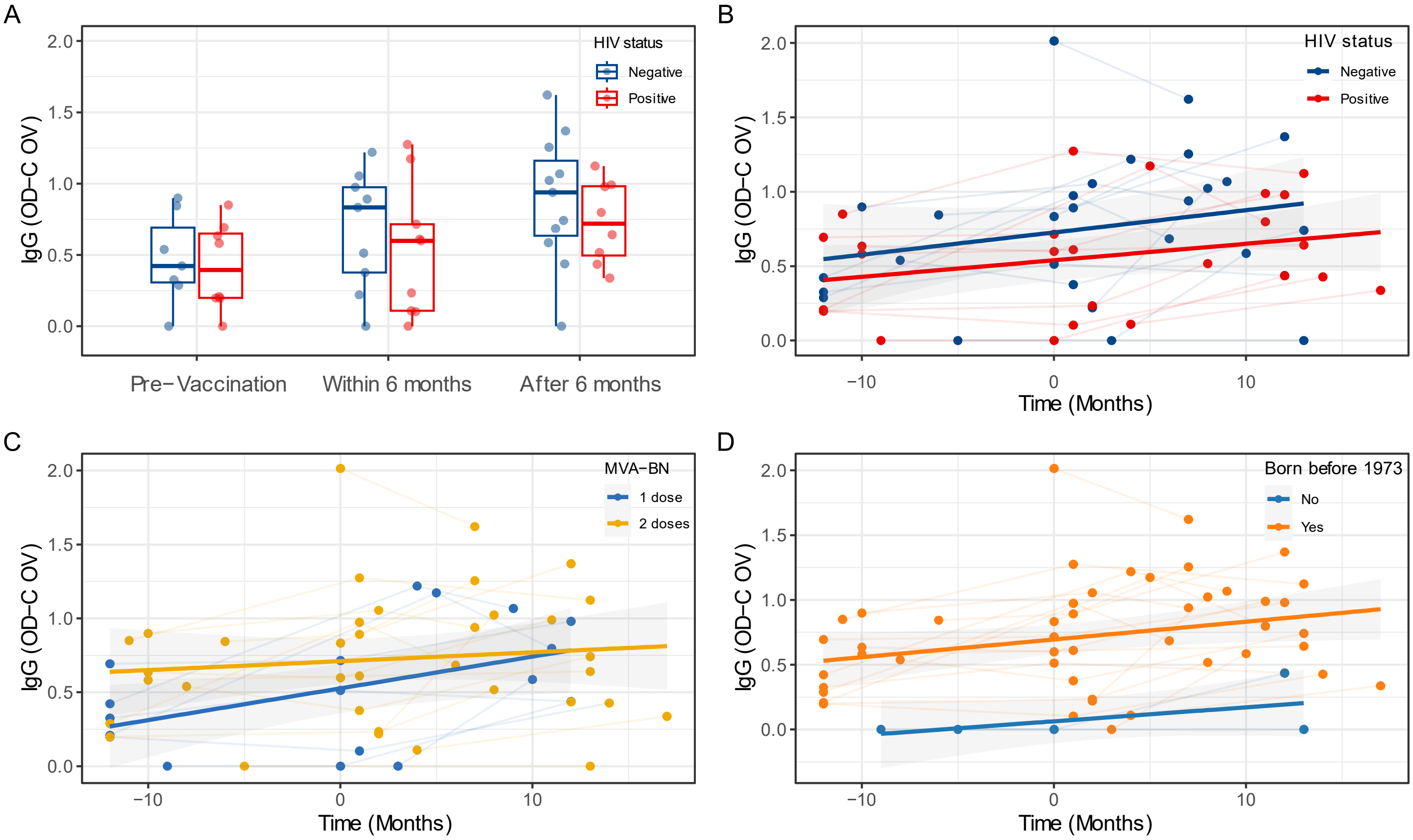


**Figure S1**. Anti-orthopoxvirus IgG levels in MVA-BN vaccinated participants. (A) IgG levels before, within 6 months of the last vaccinations, and after 6 months of the last vaccinations. P was calculated using Wilcoxon Rank Sum test but not shown (all non-significant). (B-D) Longitudinal IgG levels stratified by (B) HIV serostatus, (C) MVA-BN doses, and (D) birth before 1973. Time was calculated from the first dose of vaccinations.

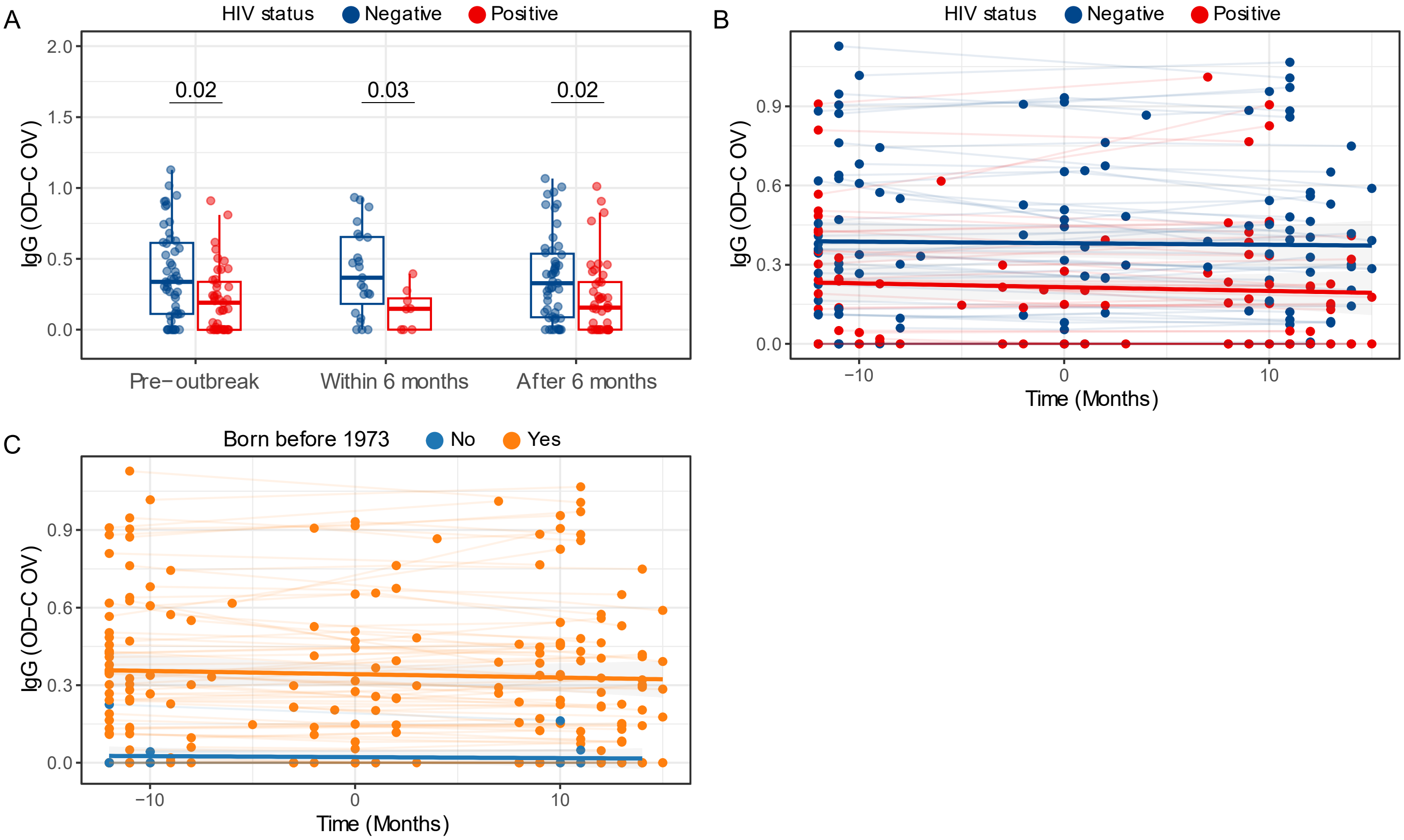


**Figure S2**. Anti-orthopoxvirus IgG levels in MVA-BN unvaccinated participants. (A) IgG levels before, within 6 months, and after 6 months of the 2022 outbreak. P was calculated using Wilcoxon Rank Sum test and adjusted for multiple comparison using the Benjamini-Hochberg method. (B-C) Longitudinal IgG levels stratified by (B) HIV serostatus, (C) birth before 1973. Time was calculated from June 1st, 2022.
